# Echocardiographic characterisation in severe Covid-19 with respiratory failure - an observational study

**DOI:** 10.1101/2023.10.27.23297666

**Authors:** Henrik Isackson, Anders Larsson, Miklos Lipcsey, Robert Frithiof, Frank A. Flachskampf, Michael Hultström

## Abstract

**Objective:** We aimed to investigate cardiac effects of severe SARS-CoV-2 and the importance of echocardiography-assessment and biomarkers.

**Methods:** This is an observational study of the first patients admitted to intensive care due to SARS-CoV-2-respiratory failure. Thirty-four underwent echocardiography of which twenty-five were included, compared to forty-four non-echo patients. Exclusion was based on absence of normofrequent sinus rhythm and/or mechanical respiratory support. Biomarkers were analysed on clinical indication.

**Results:** Mortality was higher in the echo-compared to non-echo group (44 % vs. 16%, p<0.05). Right-sided parameters were not under significant strain. Tricuspid valve regurgitation velocity indicated how increased pulmonary pressure was associated with mortality (survivors: 2.51 ± 0.01 m/s vs. non-survivors: 3.06 ± 0.11 m/s, p<0.05), before multiple comparison-correction. Setting cut-off for pulmonary hypertension to 2.8 m/s generated p<0.01 using frequency distribution testing. Cardiac markers, high sensitivity cardiac troponin I and N-terminal pro brain natriuretic peptide, and D-dimer were higher in the echocardiography group. (hs-TnI (ng/L): echo : 133 ± 45 vs. non-echo: 81.3 ± 45, p<0.01; NT-proBNP (ng/L): echo: 2959 ± 573 vs. non-echo: 1641 ± 420, p<0.001; D-dimer (mg/L): echo: 16.1 ± 3.7 vs. non-echo: 6.1 ± 1.5, p<0.01) and non-survivor group (hs-TnI (ng/L): survivors: 59.1 ± 21 vs. non-survivors: 211 ± 105, p<0.0001; NT-proBNP (ng/L): survivors: 1310 ± 314 vs. non-survivors: 4065 ± 740, p<0.0001; D-dimer (mg/L): survivors: 7.2 ± 1.5 vs. non-survivors: 17.1 ± 4.8, p<0.01). Tricuspid regurgitation velocity was positively correlated with cardiac troponin I (r=0.93, r^2^=0.74, p<0.001).

**Conclusions:** These results suggest there is no negative effect on cardiac function in critical SARS-CoV-2. Pulmonary pressure appears higher amongst non-survivors indicating pulmonary disease as the driver of mortality. Echocardiography was more commonly performed in the non-survivor group, and cardiac biomarkers as well as D-dimer was higher in the non-survivor group suggesting they carry negative prognostic values.

**Trial registration number:** This is an observational study from patients included in “Clinical trials NCT04316884”

*Strength and limitations of this study:*

- The patient cohort is recruited from consecutive patients admitted to the ICU in need of mechanical respiratory support independent of background which makes it relevant to clinical practice.
- The echocardiographic image acquisition was carried out by hospital assigned agents on clinical indication, which makes the results applicable in a clinical setting.
- Since the image acquisition was carried out on a clinical indication, the results may be skewed towards the false positive if applied to all Covid19 patients.

## Introduction

Since the severe acute respiratory syndrome corona virus 2 (SARS-CoV-2) was established as an infection transmitted amongst humans, it has been under intense scrutiny. The effects on the cardiovascular system are not completely understood.

The respiratory tract is undisputed as the main target of SARS-CoV-2, but reports of myocarditis have been published (1), with autopsy findings showing myocardial T-cell infiltration (2)(3) and SARS-CoV-2 genome in myocardial biopsies from 104 patients with suspected myocarditis (4). Cellular internalisation through the angiotensin converting enzyme 2 (ACE-2) receptor also generated initial concern that heart failure and hypertension patients treated with angiotensin (Ang) inhibitors would be at increased risk due to upregulation of ACE-2 (5) and that increased ACE-2 shedding lowers the AngI/AngII causing vasoconstriction, inflammation, and risk of thrombosis. The occurrence of venous thromboembolic disease, as well as levels of cardiac markers, troponin and natriuretic peptide, have been suggested to correlate with negative outcomes (6). Thus, patient assessment by echocardiography has attracted clinical interest.

We report from an exploratory study of the first unselected seventy-nine Covid-19 patients admitted to the intensive care unit (ICU) of a major tertiary centre. The need for echocardiography may itself raise clinical awareness of a patient’s decline which we aimed to investigate, as well as the interrelationship between clinically initiated echocardiography-results and biomarker levels.

## Methods

This is a sub-study of the single-centre, prospective observational investigation PronMed study, approved by the Swedish National Ethical Review Agency, Dnr 2017-043 (with amendments 2019-00169, 2020-01623, 2020-02719, 2020-05730, 2021-01469) and Dnr 2022-00526-01. Informed consent was obtained either from the patient or from next-of-kin if the patient was unable to receive information due to their clinical status. The Declaration of Helsinki and its subsequent revisions were followed. The protocol of the study was registered a priori (Clinical Trials ID: NCT04316884). STROBE guidelines were applied in reporting. All patients were diagnosed by polymerase chain reaction from respiratory tract swabs and had respiratory failure requiring at least high flow oxygen therapy before admission to the ICU. Patients that were investigated by echocardiography were labelled “echo” patients and those that were not investigated by echocardiography were labelled “non-echo” patients.

Echocardiographic examination was carried out on physician-deemed indication by hospital certified sonographers. Study analysis was carried out offline, independent of clinical analysis results, on TomTec^®^ software by the primary analyst and quality-controlled by a senior echocardiographer. Analyses of poor image quality were discarded at primary analyst’s discretion. Only patients with normo-frequent sinus rhythm were included for echocardiographic analysis. Pericardial effusion was quantified in the subcostal four-chamber view. For patients serially investigated by echocardiography, only the first examination was included.

Concentrations of clinically initiated biochemical cardiac blood markers during the stay in ICU, highly sensitive troponin I (hs-TnI), N-terminal pro brain natriuretic peptide (NT-proBNP), and D-dimer were compared between echo and non-echo investigated patients. Biomarker values were correlated to separate echocardiographic parameters to investigate the predictive value of such markers on cardiac function. Concentrations in the survivor and non-survivor subgroups were compared to assess the predictive value.

Frequency distribution testing between groups were carried out using Chi-squared test or Fischer’s exact test for group sizes ≤ n=5. Groups were compared using Student’s t-test for normally distributed data and Mann-Whitney’s U-test for non-normally distributed data. Correlation analysis was performed using Pearson test for normally distributed data and Spearman’s test for non-normally distributed data. Benjamini-Hochberg procedure was applied for analyses involving more than 20 parameters to reduce the risk of type 1 errors accepting a false discovery rate of 0.1, but pre-correction values are also reported and discussed. All confidence intervals are presented as standard error of the mean (SEM) unless otherwise stated. Statistical analyses and graphs utilised GraphPad Prism 5.0. Benjamini-Hochberg procedure was carried out using Microsoft Excel 2019.

## Results

### Patient selection

Out of seventy-nine patients originally included in the study, thirty-four had been assessed by echocardiography as deemed indicated by the treating physician. Out of these nine were subsequently excluded due to initial incorrect Covid-19 diagnosis, irregular heart rhythm, 3^rd^ degree AV-block, or overall unacceptable image quality, leaving twenty-five for analysis.

Forty-five patients were included in the non-echo arm, of which one was excluded due concomitant Covid-19 and diabetic ketoacidosis with no need for respiratory support, leaving forty-four patients in the non-echo group. A total of sixty-nine patients were thus included in the study (fig 1).

**Fig 1.**
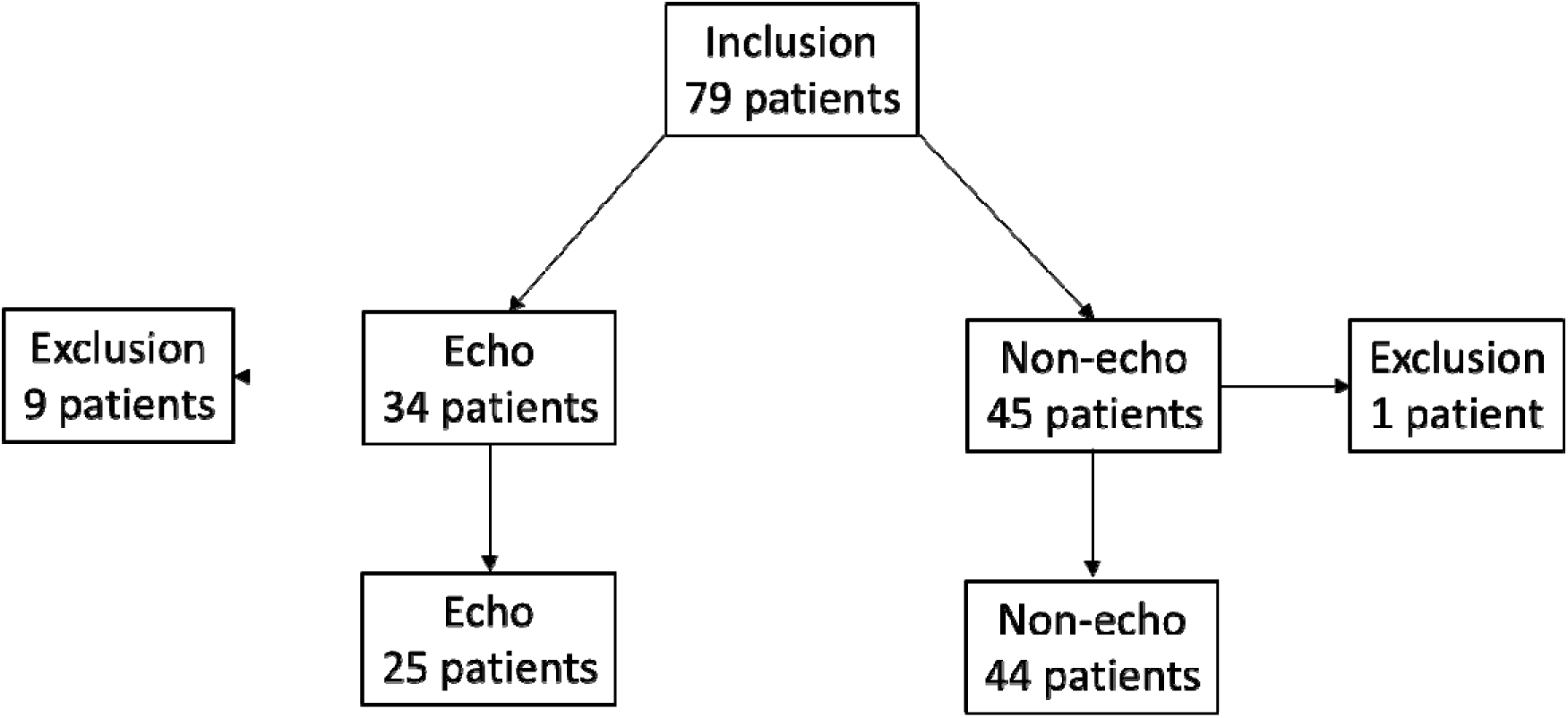
Study recruitment flow chart. Out of 79 patients included in the study, 34 were assessed by echocardiography. Out of these 9 patients were subsequently excluded (3 did not fulfil the inclusion criteria of Covid-19 infection, 4 due to irregular heart rhythm or 3^rd^ degree AV-block, and 2 due to overall unacceptable image quality), leaving 25 valid for echocardiographic analyses and inclusion. 45 patients were not assessed by echocardiography, of which one was excluded (admitted to the intensive care unit due to concomitant diabetic ketoacidosis and not Covid-19 respiratory insufficiency), leaving 44 patients not assessed by echocardiography and a total of 69 patients in the study.

### Demographic characterisation

There were more male than female patients overall (78 %), this ratio was similar in the echo group (76 %) vs the non-echo group (80 %) (p>0.05). Mean age in the echo group (64.4 ± 2.6 years) was higher than in the non-echo group (56.7 ± 2.2 years) (p<0.05). BMI in the overall cohort was in the over-weight range (28.9 ± 0.69 kg/m^2^) but there was no difference between the echo (28.8 ± 1.1 kg/m^2^) and non-echo group (29.0 ± 0.9 kg/m^2^) (p>0.05). Overall mortality in the study was 26 %, this was higher in the echo group (44 %) compared to the non-echo group (16 %) (p<0.05). Overall percentage of invasive ventilation was 55 %, also higher in the echo group (80 %) compared to in the non-echo group (41 %) (p<0.01). There was a higher presence of established ischemic heart disease (IHD) in the echo group (24 %) than in the non-echo group (4.5 %) (p<0.05) and treatment with angiotensin converting enzyme inhibitors (ACEi) or angiotensin receptor blockers (ARB) in the echo group (52 %) compared to non-echo (25 %) (p<0.05). There was no significant difference with regards to previous heart failure, hypertension, pulmonary disease, diabetes mellitus type 2, or renal failure between the echo and non-echo group (table 1).

**Table 1:**
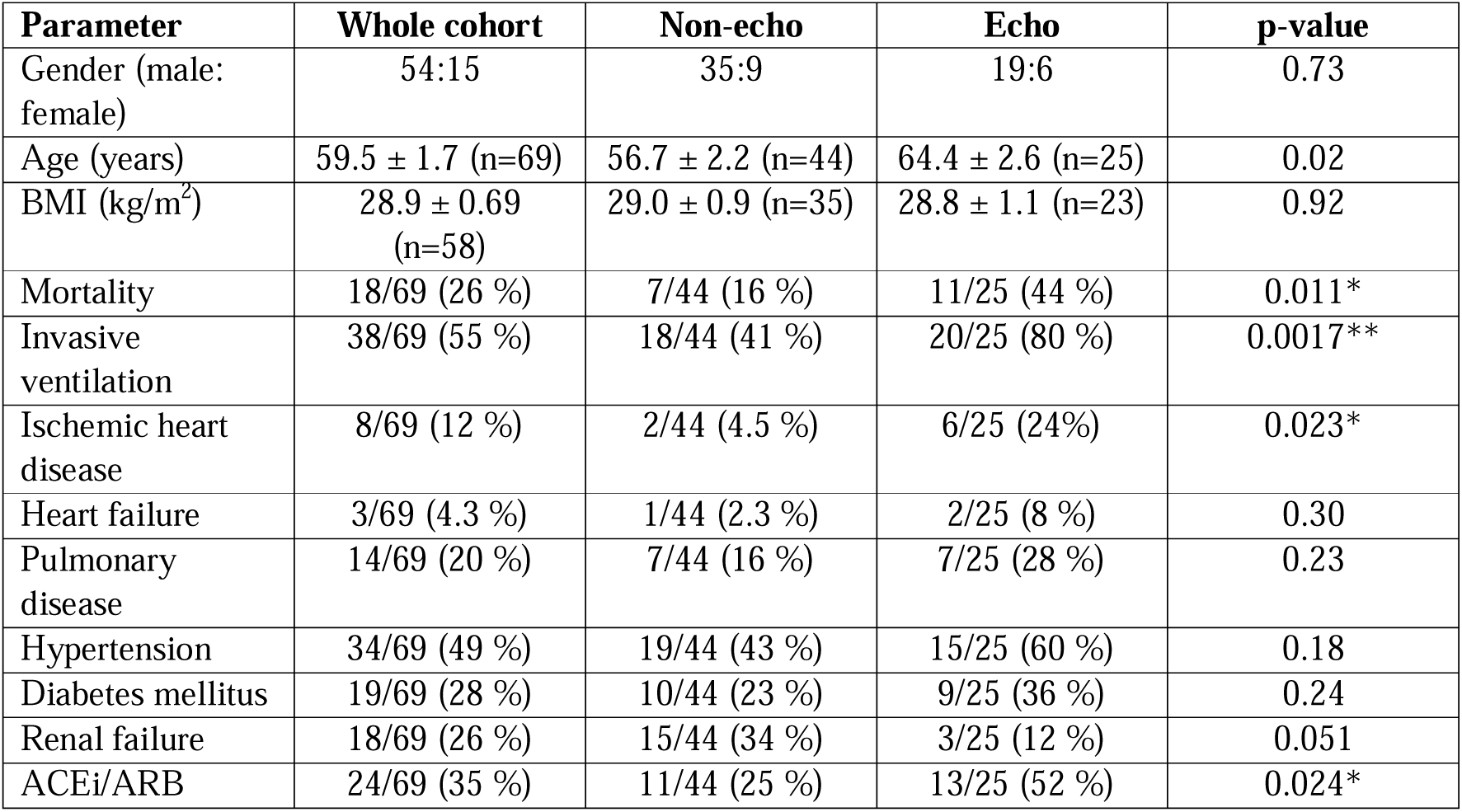
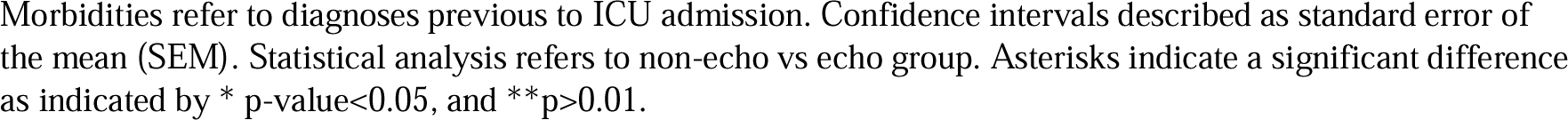
Demographic characterisation.

### Biomarkers

Admission level of hs-TnI was higher in the non-survivor group (p<0.01) and D-dimer slightly, but significantly lower in the non-survivor group (p<0.05) (table 2 a). Maximum expression levels of hs-TnI (p<0.0001), NT-proBNP (p<0.0001) and D-dimer (p<0.01) were all significantly higher in the non-survivor group (table 2 b). Maximum expression levels of hs-TnI (p<0.01), NT-proBNP (p<0.001), and D-dimer (p<0.01) was higher in the echo group compared to non-echo group (table 2 c).

**Table 2:** Relation between biomarkers, mortality and investigation by echo.

**Table 2 a:** Admission biomarker levels in survivor and non-survivor group.

| Marker | Whole cohort | Survivors | Non-survivors | p-value |
| --- | --- | --- | --- | --- |
| hs-TnI (ng/L) | 84.1 ± 40 (n=47) | 39.4 ± 15 (n=32) | 180 ± 120 (n=15) | 0.0045** |
| NT-proBNP (ng/L) | 1055 ± 276 (n=48) | 936 ± 351 (n=33) | 1318 ± 438 (n=15) | 0.25 |
| D-dimer (mg/L) | 2.58 ± 0.6 (n=63) | 2.64 ± 0.9 (n=46) | 2.42 ± 0.4 (n=17) | 0.047* |

**Table 2 b:** Maximum biomarker levels in survivor and non-survivor group.

| Marker | Whole cohort | Survivors | Non-survivors | p-value |
| --- | --- | --- | --- | --- |
| hs-TnI (ng/L) | 98.8 ± 32 (n=65) | 59.1 ± 21 (n=48) | 211 ± 105 (n=17) | <0.0001*** |
| NT-proBNP (ng/L) | 2133 ± 346 (n=67) | 1310 ± 314 (n=47) | 4065 ± 740 (n=20) | <0.0001*** |
| D-dimer (mg/L) | 9.8 ± 1.8 (n=68) | 7.2 ± 1.5 (n=50) | 17.1 ± 4.8 (n=18) | 0.0012** |

**Table 2 c:**
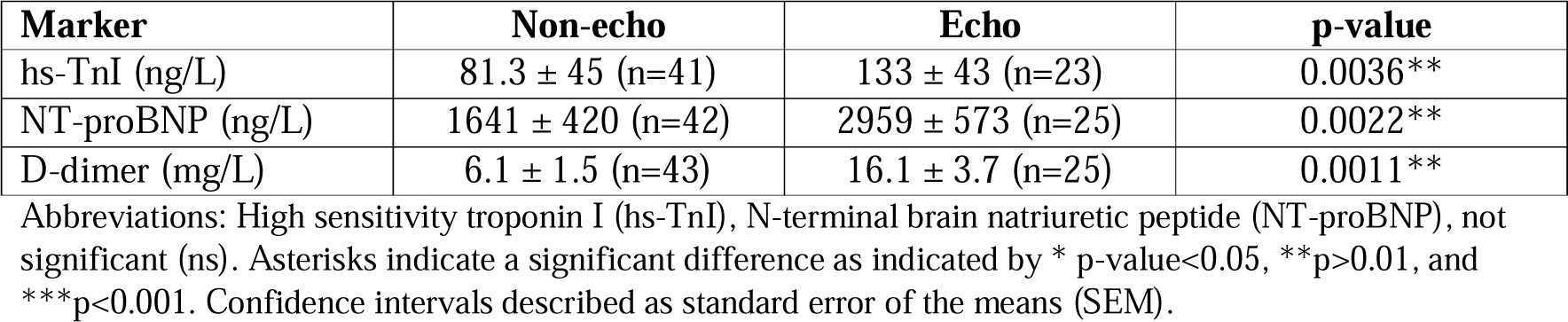
Maximum biomarker levels in the non-echo vs. echo group.

Correlation between standard echocardiographic parameters and maximum biomarker levels was investigated showing that hs-TnI was positively correlated to maximum tricuspid valve regurgitation velocity (TR V_max_) (p<0.01) (fig 2) which passed correction for multiple comparisons. A positive correlation was seen between interventricular septal diameter (IVSD) and hs-cTnI (p<0.05), early diastolic filling velocity and NT-proBNP (p<0.05), a negative correlation between NT-proBNP and right ventricle (RV) fractional area change (FAC) (p<0.05), between D-dimer and global longitudinal strain (GLS) (p<0.01) and a positive correlation between D-dimer and early diastolic filling velocity (E) (p<0.05). However, these associations were not significant when correcting for multiple comparisons. A trend towards a positive correlation between RV and RA dimension and NT-proBNP was detected that failed to reach significance (supplementary data table 1).

**Fig 2.**
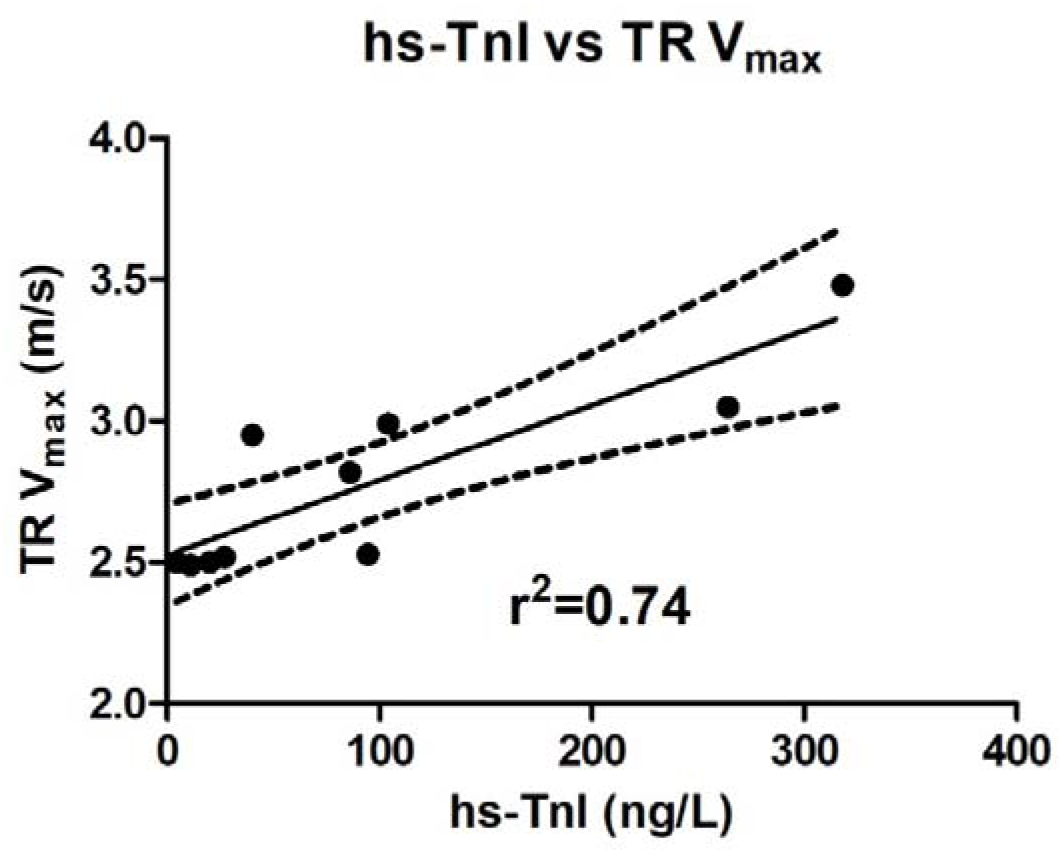
High sensitivity troponin I is correlated to maximum tricuspid valve regurgitation velocity. There was a significant correlation between maximum high sensitivity troponin I (hs-TnI) in relation to maximum tricuspid valve regurgitation velocity (TR V_max_). Linear regression fit with 95% confidence intervals and best fit. For p-and r-values, see supplementary data table 1.

### Echocardiography

In the overall echocardiographic assessment, all four heart chambers were of normal sizes. The IVSD as well as posterior wall thickness (PWT) exceeded normal values (7). Average left ventricular (LV) GLS was reduced in 17/18 patients, of which 3 had a previous diagnosis of heart failure or coronary disease. Average RV systolic function, assessed by TAPSE, RV fractional area change and RV free wall strain were within normal ranges (8). Transmitral early diastolic velocity (E) was increased but E/A ratio, septal e’, E/e’, as well as transmitral deceleration time (MV dec. time) were normal (8). IVC dimension and its respiratory variation failed to indicate clear signs of increased RA pressure (8). Average pulmonary artery acceleration time (PAAT) (9), and maximum tricuspid regurgitation velocity (TR V_max_) were normal (8). Non-survivors in ICU had a higher TR V_max_ (3.06 ± 0.11 ms) than survivors (2.51 ± 0.01 m/s) (fig 3), however not passing multiple comparison correction. Frequency distribution testing, after t-test analysis of difference between these two groups using TR Vmax >2.8 m/s as a cut-off for PH, suggested that the presence of PH was higher in the non-survivor group (p>0.01). There were no increased amounts of pericardial effusion (10). None of the scanned patients had marked valvular stenoses or regurgitations. Please see table 3 for a full list of echocardiographic values.

**Fig 3.**
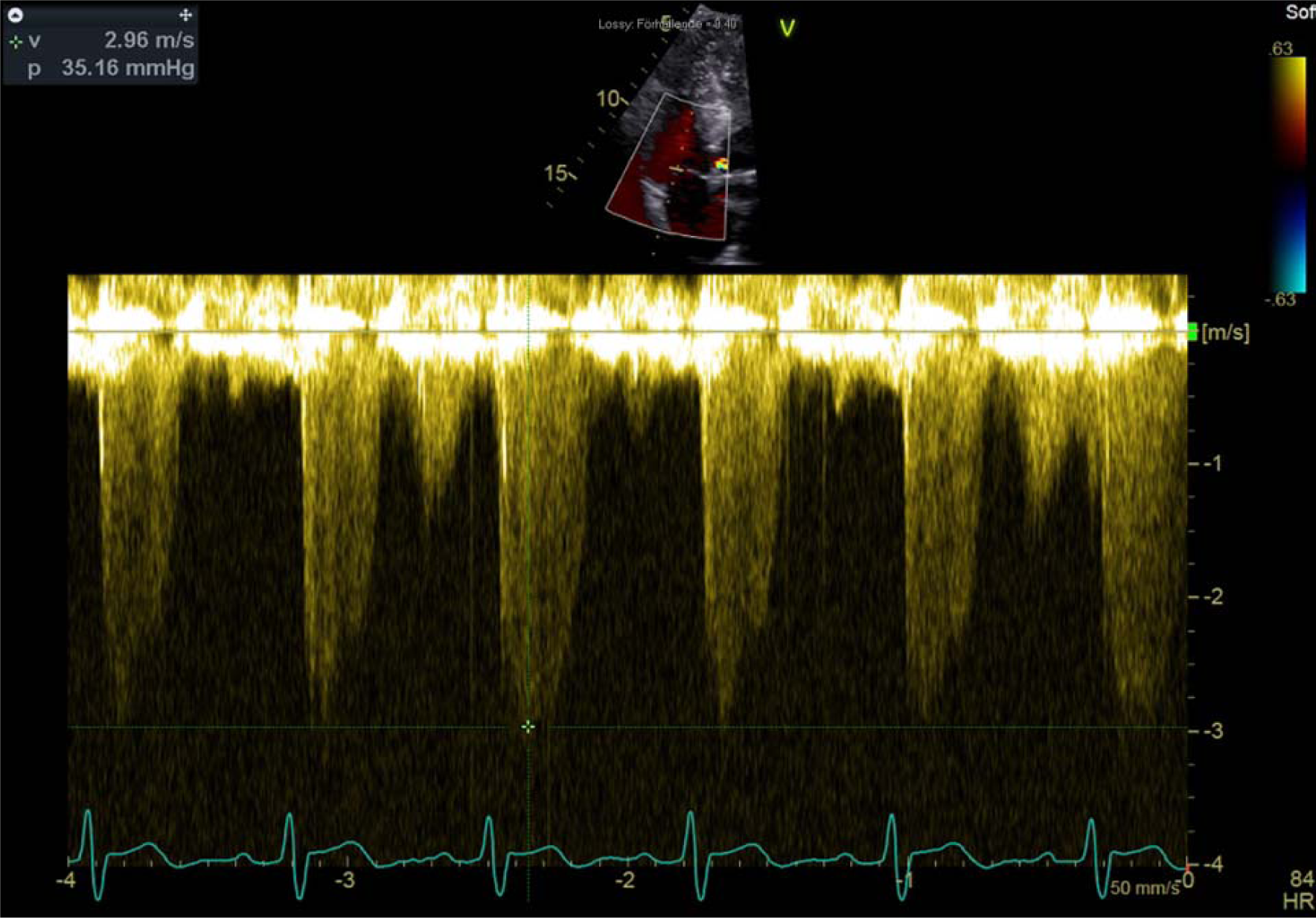
**Original continuous wave doppler trace over the tricuspid valve**. The trace derives from a patient in the non-survivor group sampled over the tricuspid valve in apical four chamber-view. TR V_max_ was the only echo-parameter that was higher amongst non-survivors than survivors which suggests pulmonary hypertension to be a negative factor in the study group. The in-picture values are derived from initial clinical scans and not used in the study.

**Table 3:** Echocardiographic parameters in the whole group and their distribution between survivor and non-survivor group.

| Parameter | Whole group | Survivors | Non-survivors | p-value (BH-significance) |
| --- | --- | --- | --- | --- |
| <u>Static measures</u> |  |  |  |  |
| IVSD (mm) | $11.4 \pm 0.5$ (n=22) | $11.7 \pm 0.58$ (n=12) | $10.9 \pm 0.88$ (n=10) | 0.42 (ns) |
| LVEDD (mm) | $47.7 \pm 1.3$ (n=22) | $45.9 \pm 1.7$ (n=12) | $49.8 \pm 1.6$ (n=10) | 0.13 (ns) |
| LVEDD/m<sup>2</sup> (mm/mm<sup>2</sup>) | $23.8 \pm 0.57$ (n=22) | $23.3 \pm 0.90$ (n=12) | $24.4 \pm 0.62$ (n=10) | 0.20 (ns) |
| PWT (mm) | $10.1 \pm 0.3$ (n=22) | $10.2 \pm 0.4$ (n=12) | $9.8 \pm 0.6$ (n=10) | 0.43 (ns) |
| LA (ml) | $47.8 \pm 3.3$ (n=20) | $44.1 \pm 3.7$ (n=11) | $52.4 \pm 5.5$ (n=9) | 0.21 (ns) |
| LA/m<sup>2</sup> (ml/m<sup>2</sup>) | $23.8 \pm 1.61$ (n=20) | $22.3 \pm 1.6$ (n=11) | $25.7 \pm 3.0$ (n=9) | 0.30 (ns) |
| RVD1 (mm) | $36.1 \pm 1.5$ (n=22) | $34.2 \pm 2.0$ (n=12) | $38.4 \pm 2.0$ (n=10) | 0.14 (ns) |
| RVD1/m<sup>2</sup> (mm/m<sup>2</sup>) | $18.0 \pm 0.61$ (n=22) | $17.4 \pm 0.85$ (n=12) | $18.7 \pm 0.86$ (n=10) | 0.31 (ns) |
| RA (ml) | 49.0 ± 6.1 (n=18) | 46.4 ± 8.2 (n=10) | 52.1 ± 9.6 (n=8) | 0.66 (ns) |
| RA/m<sup>2</sup> (ml/m<sup>2</sup>) | 24.2 ± 2.9 (n=18) | 23.7 ± 3.9 (n=10) | 24.9 ± 4.5 (n=8) | 0.85 (ns) |
| IVC diameter (mm) | 20.0 ± 0.84 (n=18) | 19.0 ± 1.4 (n=7) | 20.6 ± 1.1 (n=11) | 0.65 (ns) |
| Pericardial effusion (mm) | 0.71 ± 0.25 (n=24) | 0.97 ± 0.39 (n=13) | 0.41 ± 0.29 (n=11) | 0.31 (ns) |
| **Functional measures** |  |  |  |  |
| LVEF (%) | 54.8 ± 2.2 (n=20) | 54.1 ± 3.1 (n=11) | 55.7 ± 3.4 (n=9) | 0.68 (ns) |
| GLS (%) | -14.1 ± 0.86 (n=18) | -13.5 ± 1.3 (n=11) | -15.1 ± 1.0 (n=7) | 0.16 (ns) |
| E (m/s) | 0.77 ± 0.04 (n=20) | 0.71 ± 0.05 (n=10) | 0.83 ± 0.07 (n=10) | 0.20 (ns) |
| A (m/s) | 0.64 ± 0.03 (n=20) | 0.62 ± 0.05 (n=10) | 0.67 ± 0.05 (n=10) | 0.97 (ns) |
| E/A | 1.30 ± 0.13 (n=20) | 1.27 ± 0.23 (n=10) | 1.33 ± 0.13 (n=10) | 0.34 (ns) |
| e' medial (m/s) | 0.079 ± 0.01 (n=16) | 0.072 ± 0.01 (n=6) | 0.083 ± 0.01 (n=10) | 0.41 (ns) |
| E/e' | 10.5 ± 0.81 (n=16) | 9.72 ± 0.54 (n=6) | 10.9 ± 1.3 (n=10) | 0.64 (ns) |
| MV dec. time (ms) | 211 ± 12 (n=20) | 203 ± 21 (n=10) | 220 ± 11 (n=10) | 0.47 (ns) |
| TAPSE (mm) | 22.7 ± 1.1 (n=22) | 23.4 ± 1.5 (n=12) | 21.8 ± 1.6 (n=10) | 0.48 (ns) |
| RV FAC (%) | 45.2 ± 2.4 (n=20) | 47.9 ± 2.4 (n=10) | 42.4 ± 4.1 (n=10) | 0.26 (ns) |
| RV free wall strain (%) | -25.1 ± 2.5 (n=16) | -26.3 ± 3.5 (n=9) | -23.6 ± 3.6 (n=7) | 0.68 (ns) |
| IVC resp. variation (%) | 49.5 ± 7.3 (n=17) | 59.4 ± 11.2 (n=7) | 42.6 ± 9.5 (n=10) | 0.17 (ns) |
| PAAT (ms) | 109 ± 6.3 (n=16) | 115 ± 9.2 (n=9) | 101 ± 8.0 (n=7) | 0.30 (ns) |
| TR V<sub>max</sub> (m/s) | 2.78 ± 0.11 (n=10) | 2.51 ± 0.01 (n=5) | 3.06 ± 0.11 (n=5) | 0.012* (ns) |

Confidence intervals described as standard error of the mean (SEM). Statistical analyses refer to survivor vs. non-survivor group. BH-significance refers to Benjamini-Hochberg corrected p-value. ns=non-significant. Asterisk indicate a significant difference as indicated by \* p-value<0.05, before multivariate correction.

## Discussion

The non-invasive nature and high availability of transthoracic echocardiography makes it an attractive method of assessing critically ill patients in intensive care that are often in need of serial investigations to interpret effects of treatments on the underlying condition. During spring 2020 an increasing number of patients were admitted to intensive care due to Covid-19 respiratory failure. The lungs are the main target organ of this infection, but there have been reports of myocarditis and effects on both LV and RV function which is plausible through cellular internalisation via myocardial ACE-2 receptors and indeed, SARS-CoV-2 myocarditis including myocardial virus replication has been reported (4). In a study of 305 patients, 62 % had myocardial injury as defined by elevated cardiac troponin which was associated with LV and right RV abnormalities, higher admission rate to the ICU, and mortality (11). In 74 patients with elevated troponin, 82 % requiring mechanical ventilation, mainly RV affliction was observed and associated with elevated D-dimer but not cardiac troponins. In the same cohort LV function was described as normal to hyperdynamic (12). In another study, LV dysfunction was not associated with higher mortality nor troponin levels, and RV dysfunction only present in 3/37 patients (13). In 18 patients stratified into mild and severe Covid-19, only the severely ill showed elevated measures of troponin T (TnT), N-terminal pro brain natriuretic peptide (NT-proBNP) and D-dimer as well as increased end-diastolic LV pressures without an effect on LV ejection fraction (LVEF) or RV function (14). Right but not left sided affliction was associated with death in a cohort where 63 % of 94 patients were on mechanical ventilation support (15). In 120 patients RV function and pulmonary pressure, but not left sided parameters were identified as predictors of increased mortality (16). In 200 non-ICU-patients PH without RV affliction was associated with a worsened outcome such as death or ICU admission (17). A small study of 30 patients admitted to hospital managed to show how a smaller RV and better RV systolic function, but not signs of PH, was associated with survival (18). Post-hoc analysis of 281 serially echocardiography-investigated patients with Covid-19 ARDS, disclosed how acute cor pulmonale with RV dilatation with paradoxical septum motion, was the RV functional abnormality that was most detrimental in increasing mortality (19).

From patients admitted to the ICU at Uppsala University Hospital during the first wave of Covid-19 we attempted to investigate which parameters that are affected in this group and also how they differed between survivors and non-survivors. We also correlated standard cardiac biomarkers and D-dimer values at admission as well as the maximum values in ICU to outcome and echocardiographic parameters. In this cohort we have previously demonstrated that ECG with prior myocardial infarction pattern or acute ST-T pathology at ICU admission was associated with death, need for vasoactive treatment and higher levels of biomarkers of cardiac damage and strain (20).

The mortality in the group investigated by echocardiography was higher than those that were not assessed which suggests that these patients exhibited a physiological and biochemical status of increased cardiopulmonary strain that explains the higher mortality and should alert the treating clinician to a possible decline in patient well-being. As previously known, IHD is a main risk factor of poor prognosis (21) and would also increase the likelihood of hs-cTnI release which likely explains how these patients were more often subjected to echocardiography in our study. For overall assessment of the whole cohort, the LV systolic function as assessed by GLS was clearly lower in these patients than what to expect from standard population material (8), but the LVEF was not affected which would have been expected at those clear reductions of GLS. The image quality as well as patient cooperativity was however reduced in these patients, compared to typical patients without respiratory failure, which confounds the interpretation of this isolated parameter as evidence of myocardial damage. 3/5 patients with LVEF less than normal had a previous diagnosis of heart failure or coronary disease. Neither were there signs of increased amounts of pericardial effusion. Overall diastolic dysfunction as assessed by combined evaluation of E/e’, isolated septal diastolic movement velocity (e’), LA volume, and TR V_max_ could not be verified (22). Thus, there were no overt echocardiographic signs of myocarditis or general LV affliction in these patients. Indeed, echocardiography can be regarded as a blunt tool to detect sign of myocarditis, and oedema is more appropriately evaluated using other imaging modalities, such as cardiac magnetic resonance (CMR) (23). Myocardial inflammation as assessed by CMR in patients recently recovered from Covid-19 infection of which only a third of patients had been hospitalised, was found in as many as 60 % of patients (24). The prevalence of clinical SARS-CoV-2 myocarditis is now considered in the range of 1 % (25), making our cohort too small and the image modality too unsensitive to make any strong claims regarding milder myocarditis.

Systolic and diastolic function of the LV, as judged by LVEF, GLS, and e’, showed no association to mortality. IVSD and PWT were both slightly thicker than what to expect from reference materials, this most likely reflects a high average age in the studied cohort, and trended towards thinning in the non-survival group. These findings could suggest fluid overload and passive dilation in this group, which is supported by the increased level of NT-proBNP, thus providing no evidence of myocarditis-induced oedema. Right sided parameters indicated a trend towards association with mortality. Systolic function as assessed by TAPSE, RV FAC, and RV free wall strain, showed an overall trend towards reduction in the non-survival group and the RVD1 was also increased to wider than 35 mm in 11/22 readings of all investigated patients, being insignificantly increased to 38.4 ± 2.0 mm in the non-survivor group compared to 34.2 ± 2.0 mm in the survivor group, which may indicate increased afterload of the RV due to elevated pulmonary pressure. TR V_max_ was increased in the non-survivor group using t-test, which may suggest an elevated pressure in the pulmonary circulation. However, p-value from t-test failed to show significance after multiple analysis correction. To further statistically test the presence of PH in the non-survivor group, we used frequency distribution testing assuming a cut-off value for TR V_max_ >2.8 as indicative of PH, which generated a p-value <0.01. Average PAAT was also found to be in the intermediate range and trended towards shortening in the non-survival group, suggestive of PH. This is in good concordance with results from the larger study by Huang et al where acute cor pulmonale, often caused by increased RV afterload due to increased pulmonary artery pressure, was the most detrimental of the three investigated RV abnormalities (19). In the absence of signs of LV diastolic impairment and pulmonary embolism diagnosed in only 4/25 echo patients, it is suggested this is due to hypoxia in the pulmonary arterial bed, pulmonary vasoconstriction, and possible RV strain.

Unsurprisingly, maximum levels of hs-TnI, NT-proBNP and D-dimer were all associated with death. More interestingly, when assessing admission levels only hs-TnI was associated with mortality, with a trend for NT-proBNP. hs-TnI was also associated with TR V_max_, indicating from both biochemical and echocardiographic data that this echo parameter is of importance in predicting the outcome of Covid-19 patients in the ICU. There was a trend for NT-proBNP to correlate to increased right sided dimensions and reduced RV function, that failed to reach significance. RV FAC was negatively correlated to NT-proBNP but failed to prove significance post multiple comparison testing. This could be explained by elevated pulmonary pressure, dilation of the right sided cavities and increased release of NT-proBNP. D-dimer was negatively correlated to GLS and E, which cannot be readily explained and failed to maintain significance after multiple comparison testing (supplementary table).

The main weakness of the study in detecting cardiac affliction from SARS-CoV-2, is that not all patients underwent echocardiography, but only those where the treating physician found it indicated. This leads to a selection bias but also enabled the comparison of echo vs. non-echo patients in terms of outcome and biomarker levels. However, this selection ought to produce a cohort with more severe cardiac dysfunction, meaning that our results may over-rather than under-estimate the degree of cardiac dysfunction. In addition, the small cohort size and large set of variables increases the risk of false positive effects, again suggesting that the study should tend to over-estimate the effect of critical Covid-19 on cardiac function. This was corrected for in the analysis and reported throughout.

The ICU-setting of reduced patient cooperativity as well as respiratory support, will reduce availability of echocardiographic parameters, image quality and sensitivity of interpretation which risks failing to register significant associations. For GLS measurements 4/18 were made from 2/3 standard projections, for LVEF 2/20 were made from 1/2 standard projections and for LA volume 8/20 were made from 1/2 standard projections, since images of sufficient quality was not available.

## Conclusion

There are no convincing signs of cardiac function being systematically affected in this material of unselected Covid-19 patients admitted to intensive care, and no evidence that cardiac dysfunction is a major driver of mortality in critically ill patients with Covid-19. The need for echocardiography is more common amongst non-survivors which is new information. This may alert the treating clinician to a worse prognosis. Maximum concentrations of hs-cTnI, NT-proBNP, D-dimer and admission concentrations of hs-cTnI was significantly increased amongst the non-survivors. Amongst echocardiographic parameters, only TR V_max_ is increased in non-survivors, which indicates how the primary pulmonary affliction is the strongest determinant of the patient’s prognosis.

## Data Availability

Due to Swedish and European privacy laws individual level data cannot be shared publicly but data are available after securing appropriate ethical permissions and data transfer agreements.

## Acknowledgements

The authors thank Research nurses Joanna Wessbergh and Elin Söderman, and biobank research assistants Labolina Spång, Erik Danielsson and Philip Karlsson for their expertise in compiling the study. We are also thankful to patient advisers with regards to recruitment and answering to questions regarding participation.

## Authorship contributions

HI: Carried out the primary echo image analysis, data analysis and drafted the manuscript

AL: Provided facilities for biomarker analysis and reviewed the manuscript draft

ML: Recruited patients, and reviewed the manuscript draft

RF: Recruited patients, and reviewed the manuscript draft

FF: Controlled the echo analysis and reviewed the manuscript draft

MH: Conceptualized and designed the study, recruited patients and reviewed the manuscript draft

## Data availability

Data is available from the corresponding author on reasonable request.

---

**Supplementary table 1: Correlation of echo parameters to maximum cardiac biomarkers and d-dimer values.** (p-value, r and Benjamini-Hochberg significance when p-value<0.05).

